# The Effect of Sufentanil Administration via Bolus or Infusion on Time-to-Extubation in Cardiac Surgery: A Prospective, Randomized Clinical Trial

**DOI:** 10.64898/2026.02.05.26345493

**Authors:** Peter Ricci Pellegrino, Nicholas W. Markin, Emelind Sanchez Rodriguez, Noah A. Svec, Daniel R. McDonald, Harrison B. Wurster, Jeffrey C. Songster

**Affiliations:** Department of Anesthesiology, University of Nebraska Medical Center, Omaha, NE, USA 68198; U.S. Anesthesia Partners of Texas, P.A., 1100 Allied Dr, Plano, TX, USA 75093; Associated Anesthesiologists, P.A., Plymouth, MN, USA 55447

**Keywords:** Aged, Airway Extubation, statistics and numerical data, Analgesics, Opioid, administration & dosage, Anesthesiology, methods, Cardiac Surgical Procedures, Female, Humans, Male, Middle Aged, Pain, Postoperative, prevention & control, Prospective Study, Sufentanil, administration and dosage, Randomized Controlled Trial

## Abstract

**Background:** Intraoperative opioid administration for cardiac surgery varies greatly, with most of this variability arising from anesthesiologist and institutional practices. Anesthesiologists administer intraoperative opioids via intermittent boluses and continuous infusions. Real-world data have shown infusion administration to be a strong determinant of high intraoperative opioid exposure, but whether bolus or infusion administration of sufentanil affects post-operative outcomes is unknown.

**Methods:** We conducted a prospective, randomized, single-blind controlled trial to compare the impact of intraoperative intermittent bolus administration versus continuous infusion of sufentanil on time to extubation in adult patients undergoing nonemergent cardiac surgery with cardiopulmonary bypass at a single tertiary referral university hospital in the United States.

**Results:** The primary endpoint was the time from operating room departure to extubation in the intensive care unit. The study was terminated early for futility after an interim analysis of 50 subjects. The infusion group received statistically higher doses of intraoperative opioid. The per-protocol analysis found no statistical difference in time to extubation between the bolus group (median 2.9 hours) and infusion group (median 2.6 hours). Secondary outcomes, including post-operative pain scores, opioid consumption, ICU length of stay, and hospital stay, and adverse event rates were comparable between groups.

**Conclusions:** Intraoperative administration of sufentanil via bolus or infusion results in similar time to extubation and recovery metrics. Since continuous infusions are a strong predictor of higher total intraoperative opioid doses, protocols emphasizing administration via intermittent boluses may reduce opioid exposure without compromising recovery.

**Key Points:** *Question:* Does the method of intraoperative sufentanil administration, either by intermittent bolus or infusion, affect weaning from mechanical ventilation in the intensive care unit after cardiac surgery?

*Findings:* The method of sufentanil administration did not affect time to extubation after cardiac surgery, but the infusion group received significantly higher intraoperative opioid doses compared to the intermittent bolus group.

*Meaning:* Intermittent opioid bolus administration may reduce intraoperative opioid dosage without negatively impacting recovery after cardiac surgery.

## Introduction

Intraoperative opioid administration for cardiac surgery is highly variable^1^. Historically, opioids were considered a foundational component of intraoperative management for cardiac surgical patients due to their hemodynamic stability relative to other anesthetic agents^2^. Growing awareness of opioid-related side effects like respiratory suppression and the risk of persistent opioid use have challenged the appropriateness of this practice. A recent practice guideline from the PeriOperative Quality Initiative and the Enhanced Recovery After Surgery Cardiac Society strongly recommends against the traditional opioid-based anesthetic for cardiac surgery, albeit based on a low-to-moderate strength of evidence^3^. The gulf between traditional approaches and this contemporary consensus statement reflects the current state of clinical practice, with the large variability in intraoperative opioid dosing arising primarily from anesthesiologist and institution practices instead of patient or surgical factors^1^. Further randomized controlled trials are needed to provide strong evidence that better delineates the ideal strategies for intraoperative opioid management for cardiac surgery.

Fentanyl and sufentanil are the most commonly administered intraoperative opioids for cardiac surgery in the United States^1^. Some properties of sufentanil, including its greater potency and shorter context-sensitive half-time, may make it better-suited for contemporary fast-track surgery protocols^2^. Indeed, one study found that sufentanil allows for faster weaning from mechanical ventilation after cardiac surgery compared to fentanyl^4^.

Prior studies have focused on the optimal dose of sufentanil infusions in cardiac surgery, but most anesthesiologists rely only on intermittent bolus administration of opioids^1,5,6^. Bolus administration of opioids produces rapid increases in plasma concentration that drive desired clinical effects in highly perfused target tissue before the drug undergoes redistribution and elimination^7^. By contrast, intravenous infusions of opioids result in equilibration at a steady-state plasma and target tissue concentration and can result in more prolonged effects due to drug accumulation, and real-world data have shown that intraoperative infusions are a strong determinant of higher opioid dose^1^. No prior studies have tested whether bolus or infusion as the primary modality for intraoperative opioid administration results in superior post-operative recovery in contemporary fast-track cardiac surgery. We set out to test the effect of intermittent bolus administration of sufentanil compared to continuous sufentanil infusion on time to extubation in patients undergoing elective cardiac surgery.

## Methods

### Data and Materials Availability

The deidentified data are available from the corresponding author on reasonable request.

### Trial Design

This was a prospective, randomized trial performed at the University of Nebraska Medical Center, a tertiary referral university hospital in the Midwest region of the United States of America. The institutional review board approved the trial protocol (available as part of the online supplement). All patients provided written informed consent prior to enrollment. The trial was conducted in accordance with local regulations, Good Clinical Practice guidelines, and the principles of the Declaration of Helsinki. The study was prospectively registered (NCT04226495) on October 13, 2020. Neither patients nor the public were involved in the design, conduct, or reporting of the trial.

### Patients

Patients were eligible for screening if they were 19 to 80 years of age, scheduled for non-emergent cardiac surgical procedures on cardiopulmonary bypass including coronary artery bypass grafting, aortic valve replacement, and combined coronary artery bypass grafting with aortic valve replacement. Patients were ineligible if they had a sufentanil allergy, left ventricular ejection fraction less than or equal to 30%, moderate or severe right ventricular dysfunction, moderate or severe pulmonary dysfunction (including patients on home supplemental oxygen and/or daily bronchodilator therapy), Stage III or worse chronic kidney disease, cognitive dysfunction affecting ability to consent and candidacy for rapid extubation protocol after surgery (e.g., diagnosis of Alzheimer’s dementia and related disorders), hemodynamic instability requiring mechanical circulatory support (e.g., extracorporeal membrane oxygenation, intra-aortic balloon pump, or microaxial flow pump), or were pregnant, breastfeeding, or scheduled for repeat sternotomy or emergency surgery. Eligible patients were approached either after their evaluation in cardiac surgery clinic or during an inpatient stay prior to scheduled cardiac surgery.

### Randomization and Blinding

Informed consent was obtained from all eligible patients before study enrollment at least one day prior to their surgical procedure. We randomized patients in a 1:1 ratio to receive intraoperative sufentanil either via infusion or intermittent boluses with age-based stratification via a secure Web-based system. Age-based stratification with three strata (age < 60 years, 60-69, age > 70) was performed as this is a known determinant of time to extubation after cardiac surgery^8^. After randomization, subjects were given a unique number. Subjects and intensive care unit (ICU) staff were blinded to treatment assignments by omission. The attending cardiac anesthesiologists were unblinded but were not involved in data collection or analysis.

### Trial Intervention

Patients were administered intravenous sufentanil based on their total body weight obtained on the day of surgery. Patients in both arms were administered an induction dose of 0.4 µg/kg sufentanil rounded to the nearest 5 µg and another 0.4 µg/kg approximately 3 minutes prior to sternotomy incision. For patients in the sufentanil infusion group, an infusion of sufentanil at 0.25 µg/kg/hr was initiated after this preincision bolus and maintained until separation from cardiopulmonary bypass. After separation from cardiopulmonary bypass, the sufentanil infusion rate was reduced to 0.125 µg/kg/hr until the first sternal wire was placed, at which time the infusion was discontinued. Patients in the sufentanil bolus group received another 0.4 µg/kg sufentanil bolus 5 minutes after the initiation of cardiopulmonary bypass and a 0.2 µg/kg sufentanil bolus upon release of the aortic cross clamp. Patients randomized to sufentanil bolus treatment could be administered an additional 0.2 µg/kg bolus at the discretion of the attending anesthesiologist any time after protamine administration and before arrival to the ICU. This dosing strategy was chosen such that the dosage between the two modalities would be roughly equivalent for surgical times of approximately 4 hours.

### Anesthesia Protocol

The same anesthesia protocol was used for both trial groups. The use of other anesthetic induction agents, including midazolam, propofol, ketamine, and etomidate, was left to the discretion of the attending anesthesiologist with the caveat that the midazolam dose was limited to 5 mg. General anesthesia was maintained with isoflurane, either inhaled or via the cardiopulmonary bypass circuit, targeting an end-tidal concentration of 0.6-1.2%. Depth of anesthesia was monitored continuously through processed frontal electroencephalography in the form of the bispectral index. The protocol allowed for up to 5 mg of additional midazolam to be administered intraoperatively as an adjunct to volatile anesthetics and sufentanil. The use of intraoperative and post-operative dexmedetomidine infusions was left to the discretion of the attending anesthesiologist. All patients underwent pharmacological reversal of neuromuscular blockade prior to patient hand-over in the ICU.

### Data Collection

Baseline demographic data and coexisting conditions at baseline were collected preoperatively through the electronic medical record. Doses and times of all study medications were documented in the electronic intraoperative anesthetic record by the clinical anesthesia team per standard clinical practice. Extubation time, numeric pain rating scale at the time of extubation and in the post-extubation period, and pain medications were recorded manually and charted in the electronic medical record system per standard clinical practice by the cardiovascular ICU nursing team. Blood samples were collected for quantification of sufentanil concentration.

### Primary and Secondary Endpoints

The primary outcome was time from departure from the OR to extubation in the ICU. Prespecified secondary outcomes included numeric pain rating scale over the 24 hours after extubation, post-operative pain medication dosage in oral morphine milligram equivalents (OMMEs), ICU length of stay, and hospital length of stay. We also obtained plasma samples to quantify sufentanil concentration via an enzyme-linked immunoassay (#104919, Neogen Corporation, Lexington, KY, USA), but these data are not presented as they did not conform to known pharmacokinetic models.

### Pre-specified Per-Protocol Analysis and Safety Measures

We pre-specified intraoperative criteria that were unlikely to be related to sufentanil administration but could greatly affect an individual patient’s post-operative course as exclusions from our per-protocol analysis. These included significant intraoperative blood loss requiring high volume transfusion (defined as a total of 4 units of packed red blood cells and fresh frozen plasma), need for post-operative mechanical circulatory support (e.g., extracorporeal membrane oxygenation, intra-aortic balloon pump, or microaxial flow pump), or exclusion from the standard post-operative rapid extubation protocol used at our institution. Additionally, patients who did not receive the allocated treatment due to the clinical team’s discretion were excluded from the per-protocol analysis.

The electronic medical records of all patients were reviewed for adverse intra-operative and post-operative events by the study team. All adverse events were graded per the Common Terminology Criteria for Adverse Events Guidelines, and the likelihood for causality from the study intervention was assessed by the study team.

### Statistical Analysis

Based on one month of historical data, our prospective power analysis indicated that a sample size of 86 patients would provide the trial with 80% power to detect a clinically important 60-minute difference in time to extubation at a two-sided alpha level of 0.05. We thus planned to recruit 100 subjects with an interim data analysis after data collection was completed on 50 subjects.

The statistical analyses were performed according to both per-protocol and modified intention-to-treat approach. The per-protocol analysis is presented in the main body of the manuscript while the modified intention-to-treat analysis is presented in the online supplement. There was no missing data for the primary endpoint or for any of the secondary endpoints except for some numeric pain rating scale assessments. Statistical tests were considered significant at P values less than 0.05; two-tailed testing was used wherever possible. Continuous data that were normally distributed are presented as mean ± standard deviation, and significance testing was performed with Student’s t-test. Significance testing of numeric pain rating scale values was analyzed by repeated-measures ANOVA with sufentanil administration modality as a between-subjects factor and time as a within-subjects factor using imputation for missing values. Data that failed the Shapiro-Wilk test for normality are presented as median and interquartile range, and significance testing was performed with a non-parametric Wilcoxon rank-sum test.

Categorical data are presented as frequencies and percentages and were analyzed by chi-square testing or Fisher’s exact test when expected cell counts were less than five.

## Results

### Trial Population

From October 27, 2020, to October 28, 2021, we screened 253 patients for eligibility (Figure 1). A total of 65 patients provided written informed consent and were randomized, 32 to bolus administration of sufentanil and 33 to infusion administration of sufentanil. Of these 65 patients, 55 received their allocated treatment and none were lost to follow-up, resulting in 55 patients in the modified intention-to-treat analysis. Of these 55 patients, 48 were included in the per-protocol analysis.

**Fig. 1.**
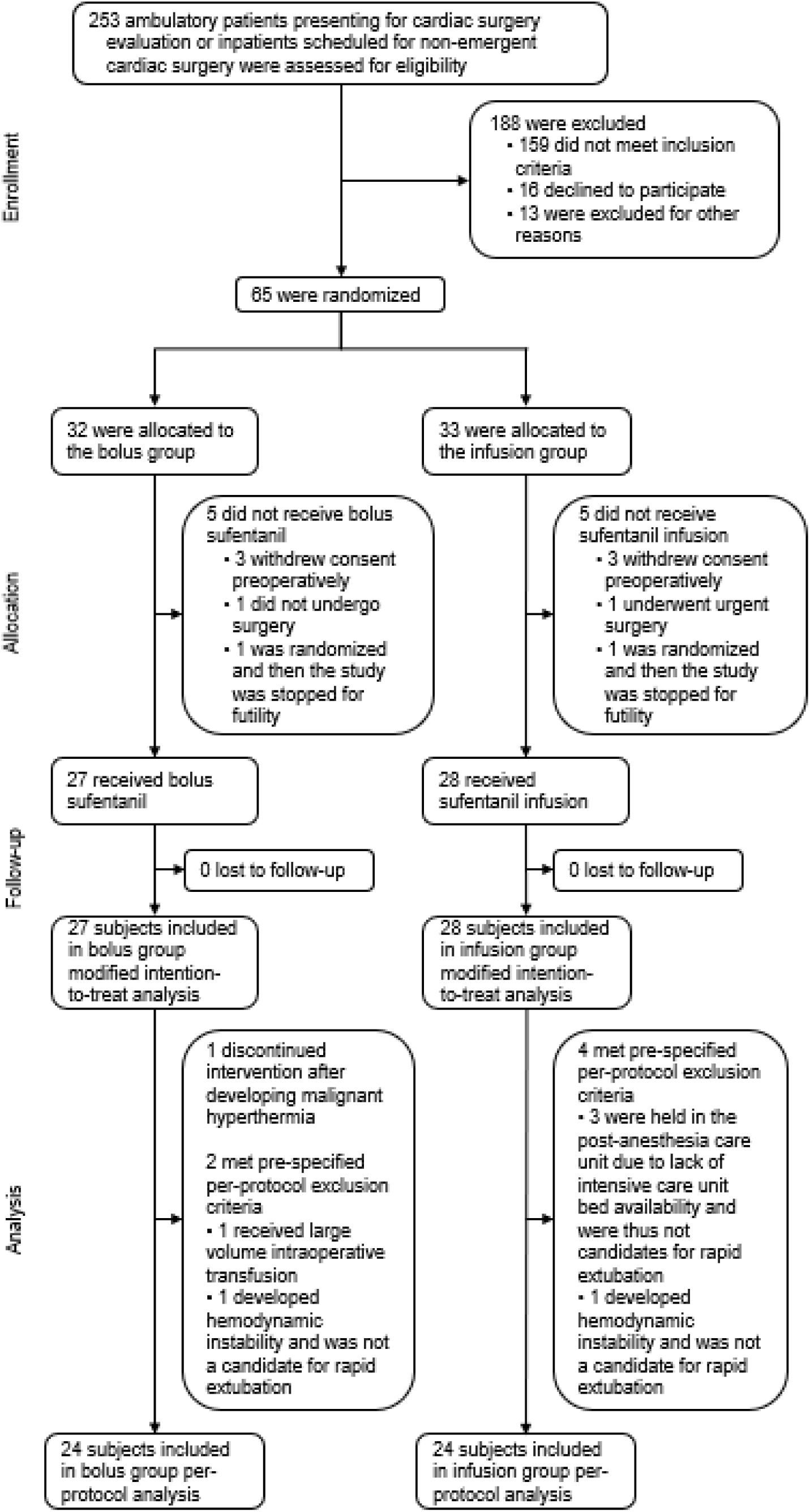
CONSORT Diagram. 253 patients were assessed for eligibility; 65 were randomized; 55 were allocated to their randomized treatment and were included in the modified intention-to-treat analysis; and 48 were included in the per-protocol analysis.

After 50 patients were recruited, the data and safety monitoring panel performed a pre-specified interim analysis, which showed the observed treatment effect size to be smaller than hypothesized. Conditional power analyses showed that even if recruitment were completed to 100 subjects and all remaining subjects demonstrated the hypothesized effect size, the primary endpoint would not approach statistical significance. Thus, the trial was stopped due to futility.

### Baseline Characteristics

At baseline, the demographic and clinical characteristics of the two groups were similar (Table 1, Table S1). Reflecting the demographics of patients undergoing coronary artery bypass grafting (CABG) and surgical aortic valve replacement (AVR) at our institution, the mean (± standard deviation) age of the subjects was 65 ± 9 years; 96% were Caucasian and 80% were male. Surgery type was highly skewed towards CABG only (80%) with 8% undergoing AVR only and 12% undergoing combined CABG and AVR. The average surgical length was 5.9 ± 1.7 hours. None of the patients had chronic outpatient opioid prescriptions.

**Table 1.**
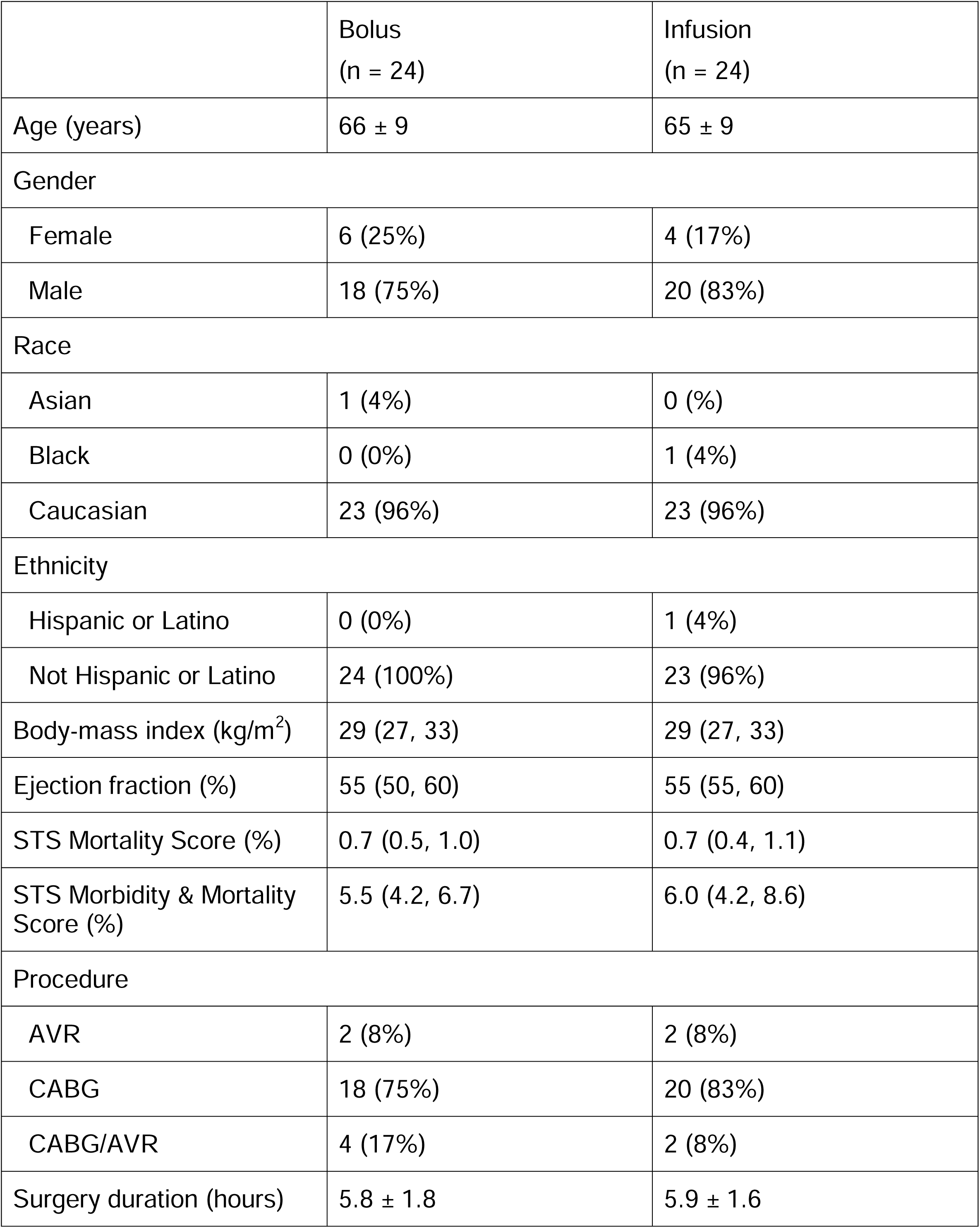
Baseline Characteristics.

|  | Bolus<br>(n = 24) | Infusion<br>(n = 24) |
| --- | --- | --- |
| Age (years) | 66 ± 9 | 65 ± 9 |
| Gender |  |  |
| Female | 6 (25%) | 4 (17%) |
| Male | 18 (75%) | 20 (83%) |
| Race |  |  |
| Asian | 1 (4%) | 0 (%) |
| Black | 0 (0%) | 1 (4%) |
| Caucasian | 23 (96%) | 23 (96%) |
| Ethnicity |  |  |
| Hispanic or Latino | 0 (0%) | 1 (4%) |
| Not Hispanic or Latino | 24 (100%) | 23 (96%) |
| Body-mass index (kg/m<sup>2</sup>) | 29 (27, 33) | 29 (27, 33) |
| Ejection fraction (%) | 55 (50, 60) | 55 (55, 60) |
| STS Mortality Score (%) | 0.7 (0.5, 1.0) | 0.7 (0.4, 1.1) |
| STS Morbidity & Mortality Score (%) | 5.5 (4.2, 6.7) | 6.0 (4.2, 8.6) |
| Procedure |  |  |
| AVR | 2 (8%) | 2 (8%) |
| CABG | 18 (75%) | 20 (83%) |
| CABG/AVR | 4 (17%) | 2 (8%) |
| Surgery duration (hours) | 5.8 ± 1.8 | 5.9 ± 1.6 |

### Trial Intervention

The trial intervention was designed such that sufentanil dose would be similar between the sufentanil bolus and infusion groups for surgeries of a duration between 3-4 hours (Figure 2A). Sufentanil dose was higher in the infusion than the bolus group (Figure 2B, Figure S1-2). As expected, sufentanil dose correlated strongly with surgery length in the infusion group and with weight in both groups (Figure 2C, Figure S1).

**Fig. 2.**
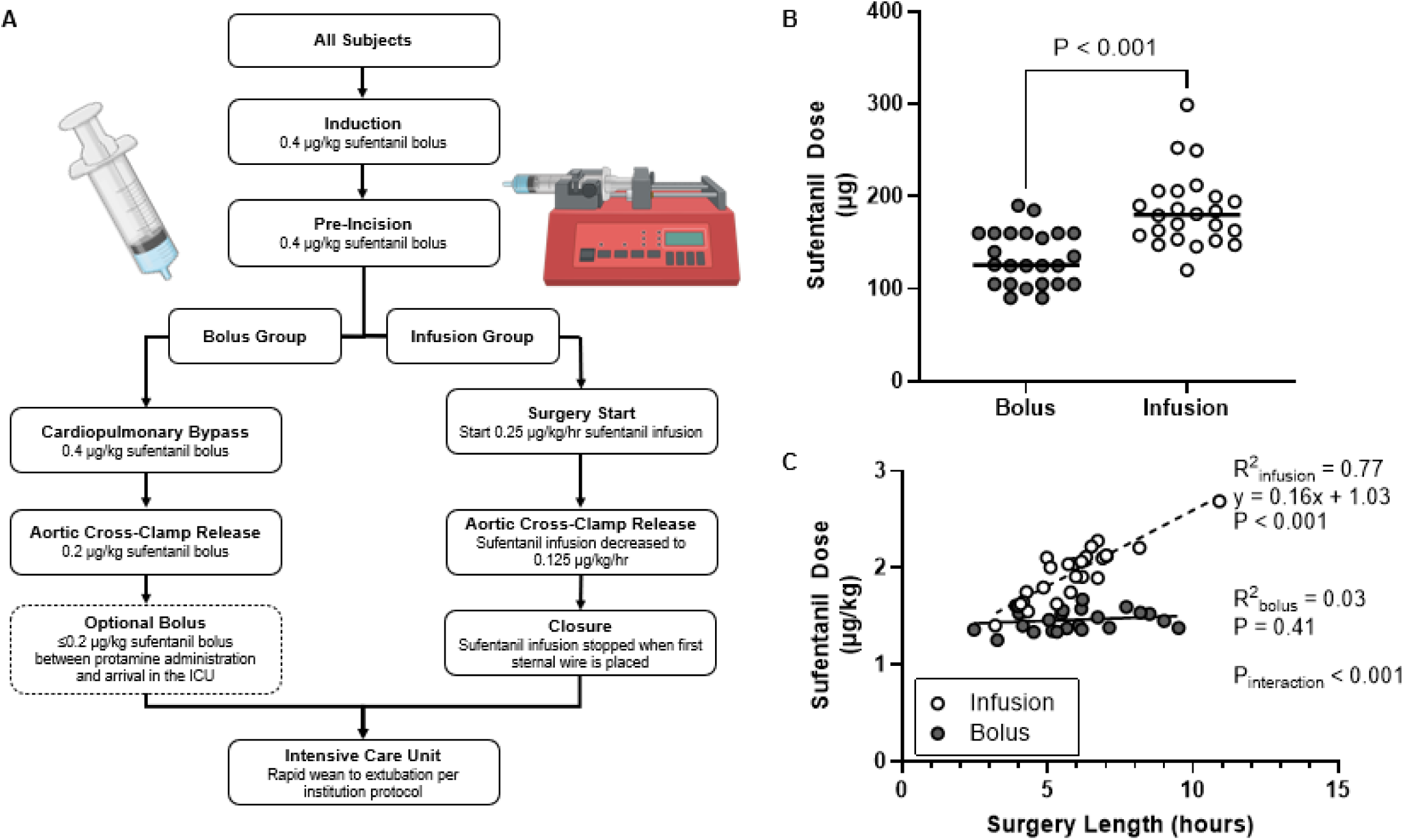
Trial intervention. (A) Protocolized intervention flow diagram showing dosing strategies and administration times for the bolus and infusion groups. (B) Total sufentanil dose was higher in the infusion group than the bolus group. (C) Weight-based sufentanil dose was higher in the infusion group and correlated strongly with surgery duration for the infusion group only.

### Time to Extubation

The primary endpoint for this randomized clinical trial was the time from operating room departure to extubation in the ICU. A standardized rapid extubation protocol was used for all patients included in the per-protocol analysis (Figure 3A). The median time to extubation was 2.9 hours in the bolus group and 2.6 hours in the infusion group, which was not statistically different (Figure 3B). The modified intention-to-treat analysis, which included patients that were not eligible for the rapid extubation protocol, showed a median time to extubation of 3.3 hours for the bolus group and 3.2 hours for the infusion group (Figure S3). Subgroup analysis did not identify any significant interactions between sufentanil infusion and baseline characteristics (Figure 3C).

**Fig. 3.**
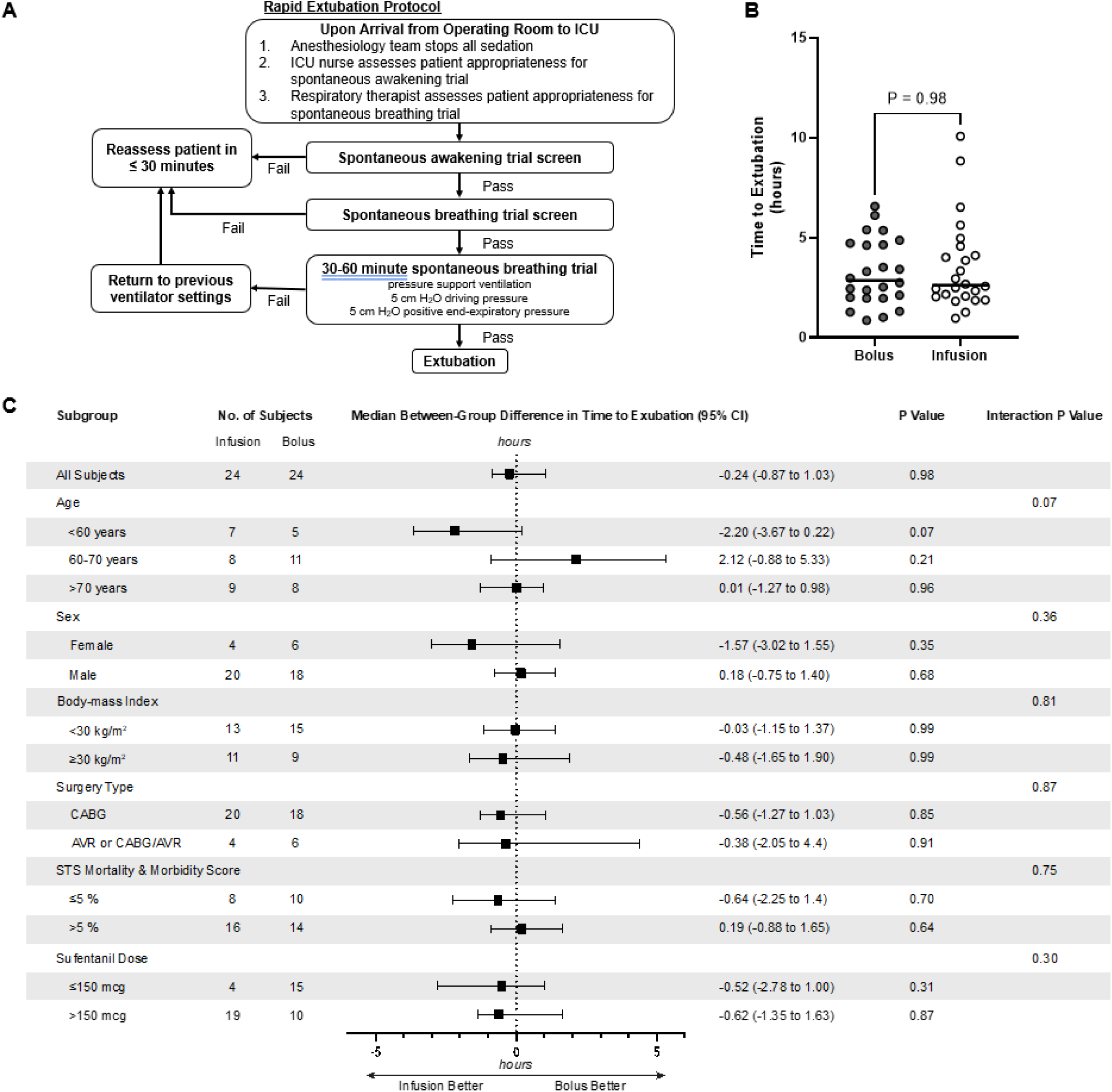
Time to Extubation. (A) The flow diagram for the rapid weaning of mechanical ventilation used for all subjects in the per-protocol analysis. (B) Bolus or infusion administration of sufentanil did not affect time to extubation. (C) Forest plot showing our post-hoc subgroup analysis, which did not identify any statistically significant effects.

### Secondary Endpoints

We also studied the effects of our trial intervention on post-operative pain, opioid use, ICU length of stay, and hospital length of stay for subjects in the trial (Figure 4, Figure S4). For both groups, the numeric pain rating scale decreased over the 24 hours after extubation, but the method of intraoperative sufentanil administration did not affect pain scores or this trend. Both groups also received similar amounts of opioid analgesics during their ICU stay (41 OMMEs for the bolus group; 34 OMMEs for the infusion group, P = 0.92). There were no statistical differences in ICU stay (P = 0.11) or post-operative hospital stay (P = 0.23) between the bolus and infusion groups.

**Fig. 4.**
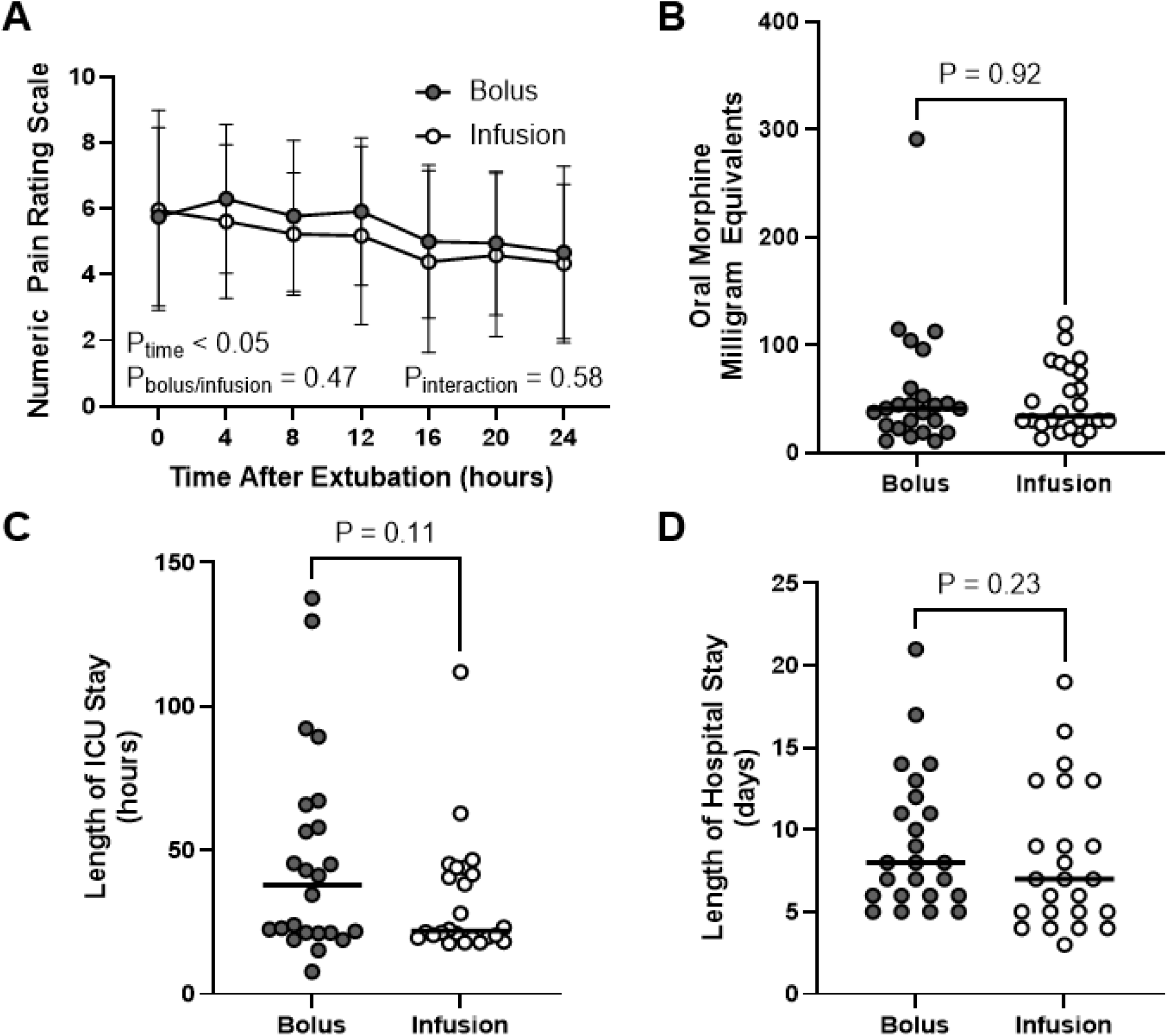
Secondary Endpoints. Sufentanil administration by bolus and infusion resulted in similar post-operative outcomes with respect to (A) patient-reported analgesia, (B) opioid requirements, (C) length of post-operative ICU stay, and (D) length of post-operative hospital stay.

### Adverse Events / Safety Outcomes

There were no statistically significant differences between vasopressor and inotrope requirements at the time of departure from the operating room between the bolus and infusion groups (Table 2, Table S2). There was a trend toward a higher vasoactive-inotropic score in the bolus group in the per-protocol analysis (P = 0.052) that was less pronounced in the modified intention-to-treat analysis (P = 0.11), suggesting this to be due to chance.

**Table 2.**
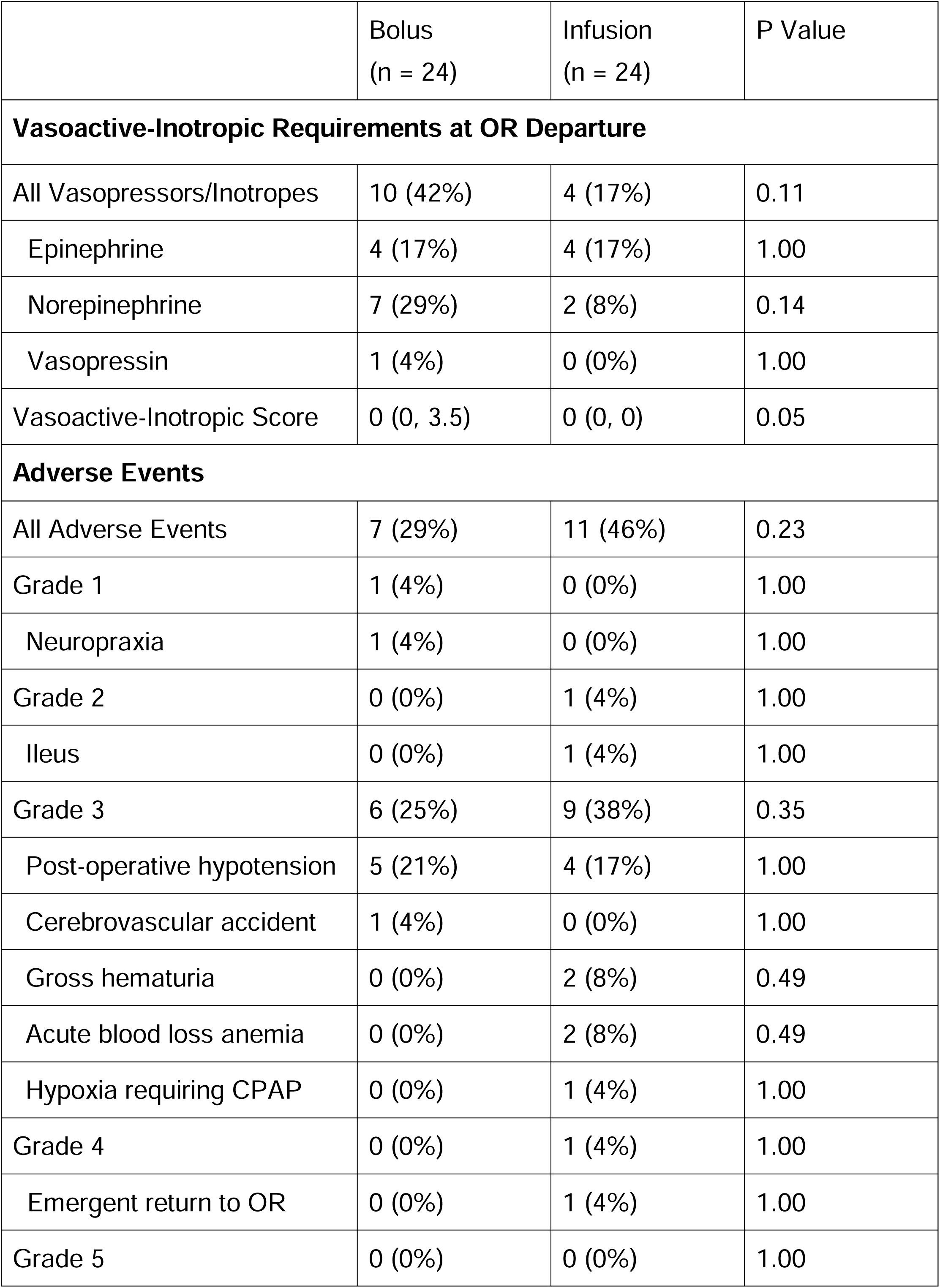
Safety Outcomes.

A total of 38% of patients in the per-protocol analysis and 41% of patients in the modified intention-to-treat analysis experienced adverse events. The rates for adverse events were similar between groups. There were only two events that were possibly related to the trial intervention. In the per-protocol sufentanil infusion group, one patient experienced a clinically significant post-operative ileus which was managed conservatively. In the modified intention-to-treat infusion group, one patient demonstrated patient-ventilator dyssynchrony with resultant profound hypotension that required neuromuscular blockade, making the patient no longer a candidate for the rapid extubation protocol. Overall, the safety profiles of bolus and infusion administration of sufentanil were similar.

## Discussion

In this single-center randomized controlled trial, we found that administration of sufentanil via bolus and infusion resulted in similar time to extubation for patients undergoing routine cardiac surgery with CPB. We did not find any differences with respect to patient-reported post-operative analgesia, post-operative opioid requirements, or time to discharge from the ICU and hospital. The immediate post-operative vasoactive and inotropic infusion requirements and adverse event rates of both administration strategies were comparable.

Despite designing the interventions such that the total intraoperative doses of sufentanil would be roughly equivalent for the two groups, the infusion group received both statistically and clinically significantly more intraoperative opioid. This stemmed from surgical times being longer than anticipated. Interestingly, this recapitulates real-world data from a recent multicenter study of practice which found that intraoperative infusions were a powerful predictor of higher opioid administration in cardiac surgery^1^. For context, using a conversion factor of 1:11, the mean dose in fentanyl equivalents for the bolus group was 1381 fentanyl µg equivalents, slightly higher than the median of 1000 fentanyl µg equivalents in this 30-hospital study of practice patterns. The median dose in the infusion group, however, was 1981 fentanyl µg equivalents, corresponding to 22 fentanyl µg equivalents per kg body mass which exceeds the 20 µg/kg cut-off that has been used to define high-opioid cardiac anesthesia^9^. Limiting opioid administration to intermittent bolus dosing can reduce intraoperative opioid exposure, which may in turn decrease the risk of opioid-related side effects.

This same real-world multicenter study of practice found that over 60% of the variability in intraoperative opioid administration was explained by anesthesiologist and institution^1^. As one would expect, the profound variability between providers present in real-world clinical practice was greatly limited by the protocolized nature of the trial intervention, with weight and surgery length explaining the majority of the variability in opioid administration. Thus, our study highlights the opportunity for protocolized approaches, like those used for enhanced recovery after surgery, to bring patient and surgical factors to the forefront, standardize clinical practice, and greatly reduce the heterogeneity in opioid administration for cardiac surgical patients.

Opioids provide potent analgesia and blunt the surgical stress response but can produce respiratory depression, sedation, nausea, vomiting, constipation, hyperalgesia, persistent use, and death^2,10^. In the setting of mounting appreciation of these adverse effects, intraoperative opioid administration has decreased^1^. The contemporary opioid-sparing approach does more than simply reduce intraoperative opioid; instead, non-opioid interventions like acetaminophen, cyclooxygenase-2 inhibitors, Na_V_1.8 inhibitors, gabapentinoids, dexmedetomidine, and local/regional anesthesia are incorporated to provide some of the benefits of opioids while reducing opioid-related side effects. This is a domain of evolving clinical practice and active investigation with key questions surrounding the impacts, costs, risks, and benefits of these alternative approaches.

Approximately 5-10% of patients develop persistent opioid use after cardiac surgery^11,12^. While strong data links perioperative and particularly post-operative opioid use to persistent opioid use, the evidence that intraoperative opioids drive persistent opioid use is mixed^11^. On one hand, a single-center cohort study of noncardiac surgical patients found that higher doses of intraoperative opioids were associated with decreased postoperative pain and reduced persistent opioid use^13^. On the other hand, randomized controlled trials of cardiac surgical patients have found that lower target concentrations of sufentanil reduce post-operative pain and facilitate rapid extubation after cardiac surgery^5,6^, and a recent meta-analysis of cardiac surgery found that opioid-sparing approaches reduce post-operative pain and opioid use^14^. Other studies have found no signal for benefit or harm with opioid-sparing cardiac anesthetic approaches^9,15^. The heterogeneity in these results reflects not only fundamental differences in the studies themselves but also differing and rapidly changing standards of care.

The strengths of this study include that it is a prospective, randomized controlled trial addressing a relevant clinical question and significant gap in the existing literature. The study also boasts excellent protocol adherence and very little missing data. The study also has limitations. The fact that this study was stopped early for futility limited our ability to detect small differences in secondary outcomes, although unpowered secondary outcomes should always be interpreted with caution. As a single-center study of patients who underwent procedures with CPB, institutional factors may also limit the generalizability of these findings to other practices. Since our patients were extubated in the ICU a median duration of 2.7 hours after departing from the operating room, these results likely do not apply to practices where extubation is commonly performed in the operating room after cardiac surgery without CPB^16^.

In summary, administration of intraoperative sufentanil via either intermittent bolus or infusion results in similar time to extubation and other key post-operative outcomes for adult patients undergoing routine cardiac surgical procedures. Transitioning away from intraoperative opioid infusions represents a simple way to decrease intraoperative opioid administration. Further studies linking intraoperative opioid administration to important post-operative outcomes like chronic pain and persistent opioid use would help characterize the value of opioid-sparing approaches to cardiac surgery.

## Data Availability

All data produced in the present study are available upon reasonable request to the authors.

## Acknowledgments

The authors would like to thank Julie Hoffman, Lace Sindt, Valerie Shostrom, Gurudutt Pendyala, and Sean Avedissian for their technical assistance.

## Supplemental Material

Figures S1-S4

Tables S1-S2

Study Protocol

