## Supplementary material for "The Effect of Sufentanil Administration via Bolus or Infusion on Time-to-Extubation in Cardiac Surgery: A Prospective, Randomized Clinical Trial": Online Supplement

#### ONLINE SUPPLEMENT FILE

**Authors:** Peter Ricci Pellegrino[1], Nicholas W. Markin[1], Emelind Sanchez Rodriguez[1], Noah A. Svec[1], Daniel R. McDonald[2], Harrison B. Wurster[3], Jeffrey C. Songster[1]

**Affiliations:**

[1]Department of Anesthesiology, University of Nebraska Medical Center, Omaha, NE, USA 68198.

[2]U.S. Anesthesia Partners of Texas, P.A., 1100 Allied Dr, Plano, TX, USA 75093.

[3]Associated Anesthesiologists P.A., Plymouth, MN, USA 55447.

*; 984455 Nebraska Medical Center, Omaha, NE 68198-

**Supplementary Figure 1. Sufentanil dose as a function of weight by per-protocol analysis.** (A) Weight-normalized sufentanil dosage was higher in the infusion group than the infusion group. (B) As expected, sufentanil dosage correlated strongly with weight for both the bolus and the infusion groups.

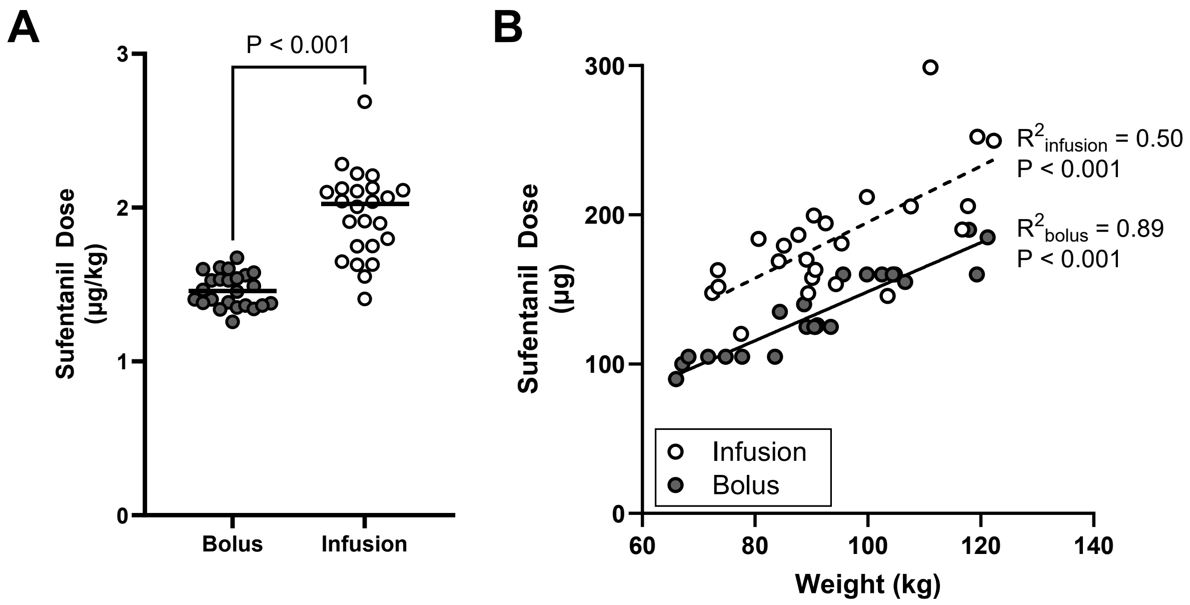

**Supplementary Figure 2. Sufentanil dose for the modified intention-to-treat analysis.** Sufentanil dosage was higher in the infusion group than the bolus group for the modified intention-to-treat subjects whether expressed as (A) total micrograms or (B) normalized by body mass.

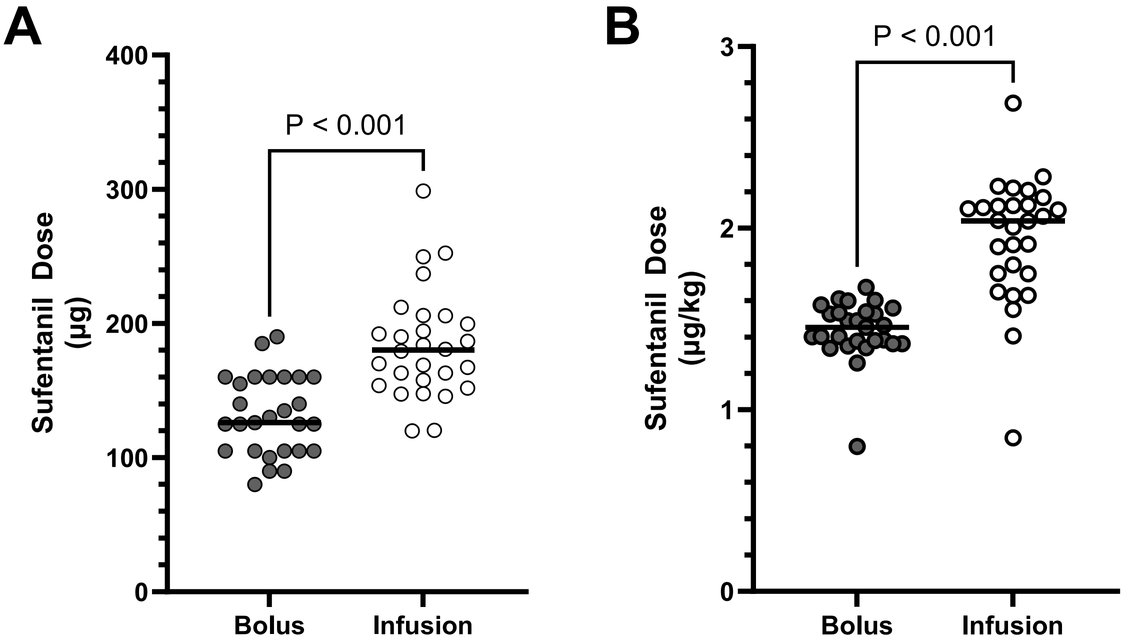

**Supplementary Figure 3. Time to extubation by modified intention-to-treat analysis.** The trial intervention did not affect time to extubation, the primary endpoint for the study, when analyzed by the modified intention-to-treat principle.

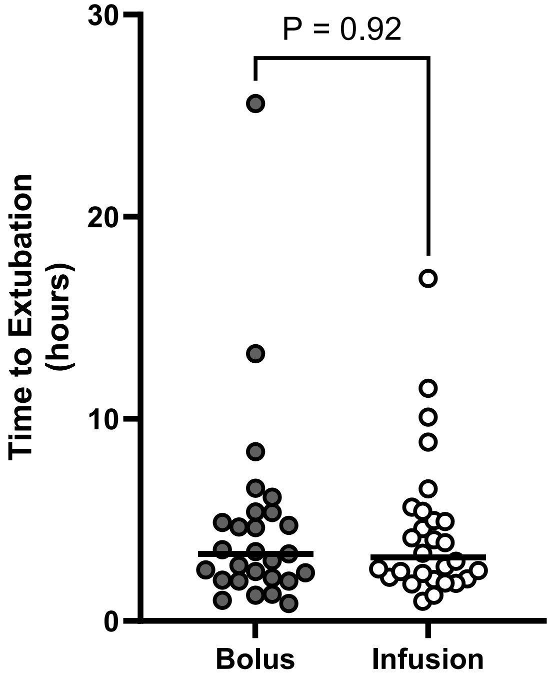

**Supplementary Figure 4. Secondary endpoints by modified intention-to-treat analysis.** When analyzed by the modified intention-to-treat principle, the trial intervention did produce statistically significant differences in (A) patient-reported analgesia, (B) opioid requirements, (C) length of post-operative ICU stay, and (D) length of post-operative hospital stay.

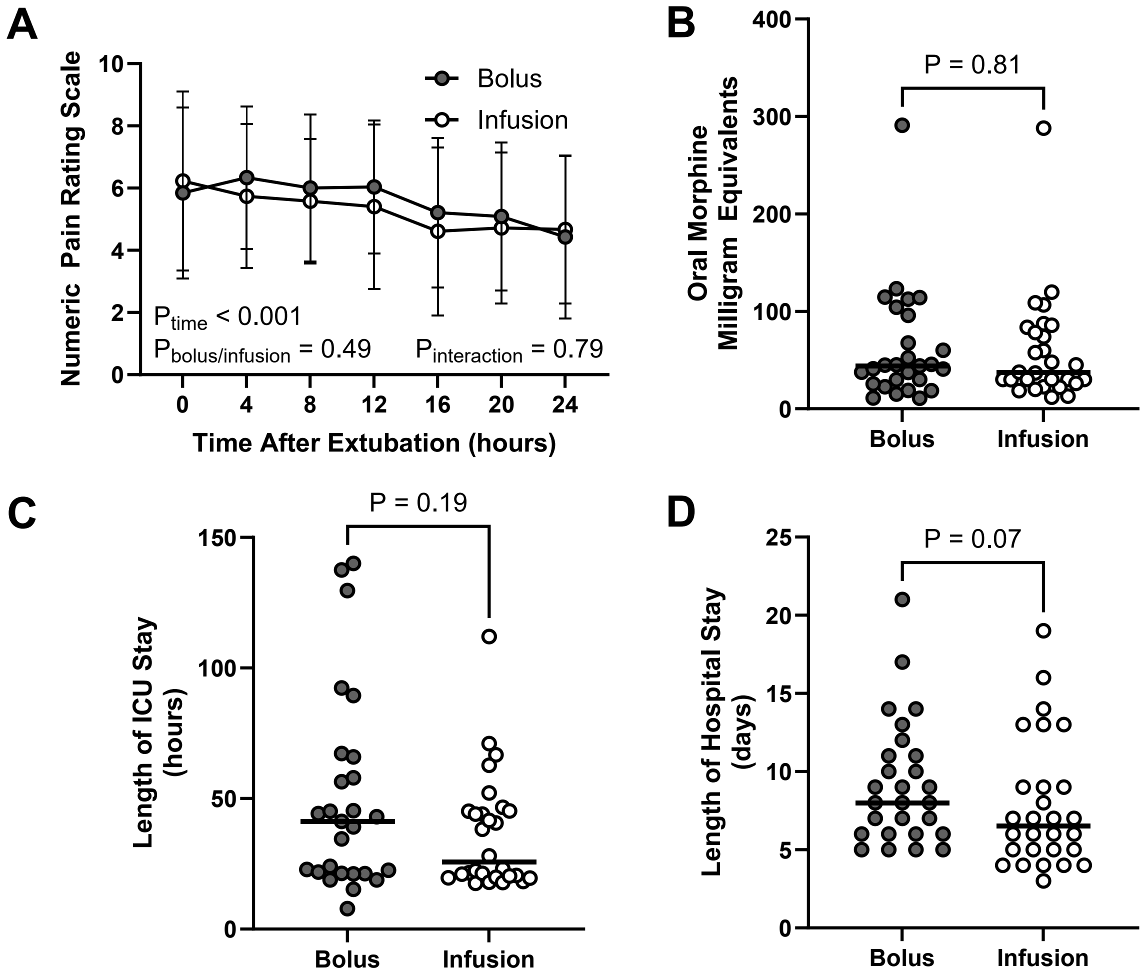

**Supplementary Table 1. Baseline characteristics per modified intention-to-treat analysis.**

| | Bolus (n = 27) | Infusion (n = 28) |
|---|---|---|
| Age (years) | 66 ± 8 | 64 ± 9 |
| Gender | | |
| Female | 6 (22%) | 5 (18%) |
| Male | 21 (78%) | 23 (82%) |
| Race | | |
| Asian | 1 (4%) | 0 (%) |
| Black | 0 (0%) | 1 (4%) |
| Caucasian | 26 (96%) | 27 (96%) |
| Ethnicity | | |
| Hispanic or Latino | 0 (0%) | 1 (4%) |
| Not Hispanic or Latino | 27 (100%) | 27 (96%) |
| Body-mass index (kg/m$^2$) | 29 (27, 33) | 30 (27, 35) |
| Ejection fraction (%) | 55 (50, 60) | 55 (53, 60) |
| STS Mortality Score (%) | 0.7 (0.5, 1.0) | 0.7 (0.5, 1.2) |
| STS Morbidity & Mortality Score (%) | 5.5 (4.4, 7.1) | 6.3 (4.6, 8.8) |
| Procedure (%) | | |
| AVR | 2 (7%) | 2 (7%) |
| CABG | 21 (78%) | 22 (79%) |
| CABG/AVR | 4 (15%) | 4 (14%) |
| Surgery duration (hours) | 6.0 ± 2.2 | 5.7 ± 1.9 |

**Supplementary Table 2. Safety outcomes per modified intention-to-treat analysis.**

| | Bolus (n = 27) | Infusion (n = 28) | P Value |
|---|---|---|---|
| **Vasoactive-Inotropic Requirements at OR Departure** | | | |
| Vasopressors/Inotropes | 11 (41%) | 6 (21%) | 0.15 |
| Epinephrine | 4 (15%) | 6 (21%) | 0.73 |
| Norepinephrine | 8 (30%) | 3 (11%) | 0.10 |
| Vasopressin | 1 (4%) | 0 (0%) | 0.49 |
| Vasoactive-Inotropic Score | 0 (0, 3.75) | 0 (0, 0) | 0.11 |
| **Adverse Events** | | | |
| All Adverse Events | 9 (33%) | 14 (50%) | 0.21 |
| Grade 1 | 1 (4%) | 0 (0%) | 0.49 |
| Neuropraxia | 1 (4%) | 0 (0%) | 0.49 |
| Grade 2 | 0 (0%) | 1 (4%) | 1.00 |
| Ileus | 0 (0%) | 1 (4%) | 1.00 |
| Grade 3 | 7 (26%) | 10 (36%) | 0.43 |
| Post-operative hypotension | 6 (22%) | 5 (18%) | 0.69 |
| Cerebrovascular accident | 1 (4%) | 0 (0%) | 0.49 |
| Gross hematuria | 0 (0%) | 2 (7%) | 0.49 |
| Acute blood loss anemia | 0 (0%) | 2 (7%) | 0.49 |
| Hypoxia requiring CPAP | 0 (0%) | 1 (4%) | 1.00 |
| Grade 4 | 1 (4%) | 3 (11%) | 0.61 |
| Patient-ventilator dyssynchrony requiring neuromuscular blockade | 0 (0%) | 1 (4%) | 1.00 |
| Post-CPB biventricular dysfunction | 0 (0%) | 1 (4%) | 1.00 |
| Malignant hyperthermia | 1 (4%) | 0 (0%) | 0.49 |
| Emergent return to OR | 0 (0%) | 1 (4%) | 1.00 |
| Grade 5 | 0 (0%) | 0 (0%) | 1.00 |

**Clinical Trial Template**

**FULL PROTOCOL TITLE**

Sufentanil Infusion vs. Sufentanil Bolus and Time to Extubation during Routine Cardiac Surgery

**Study Chairman or Principal Investigator:**

**Jeffrey Songster, MD**

Assistant Professor, Department of Anesthesiology

University of Nebraska Medical Center

**Supported by:**

Department of Anesthesiology
University of Nebraska Medical Center

**Study Intervention Provided by:**

*No support from pharmaceutical company or device manufacture.*

**NCT Number:**

NCT # 04226495

**Version 1** *(Please change version number with each amendment)*
**Month Day, Year**

**TABLE OF CONTENTS**

Page

**PRÉCIS ........ iv**

Study Title ........ iv

Objectives ........ iv

Design and Outcomes ........ iv

Interventions and Duration ........ iv

Sample Size and Population ........ iv

9.2 Interim analyses and Stopping Rules ........ 10

9.3 Outcomes ........ 16

11 PARTICIPANT RIGHTS AND CONFIDENTIALITY ........ 11

11.1 Informed Consent Forms ........ 11

12 REFERENCES ........ 11.

13 SUPPLEMENTS/APPENDICES ........ 11

**I. Procedures Schedule**

**II. Informed Consent Form Template**

**III. Other** *(add as many appendices as necessary)*

#### PRÉCIS

*This section should provide a brief protocol summary of approximately 1-2 pages.*

##### Study Title

Sufentanil Infusion vs. Sufentanil Bolus and Time to Extubation during Routine Cardiac Surgery

##### Hypothesis

We hypothesize that routine cardiac surgery patients receiving sufentanil infusion will have a lower sufentanil concentration at the end of surgery and have a shorter time to extubation than patients receiving sufentanil bolus dosing even with similar total doses of sufentanil.

##### Objectives

Primary Outcome: Time the subject becomes extubated from stop data collection time point within the EMR.

Secondary Outcome: Plasma Sufentanil concentrations, Sufentanil pharmacokinetics, 24 hour post-operative pain scores, 24 hour opioid requirements in morphine equivalents, vasopressors and inotropes usage, ICU length of stay, Hospital length of stay, early re-intubation rate.

##### Design and Outcomes

Randomized single blind controlled clinical trial to test the benefit of sufentanil infusion over bolus dosing to reduce time to extubation and reduced length of ICU stay.

Interventions and Duration:
Intervention period: intra-operative sufentanil infusion vs intermittent bolus dosing the intervention will stop at the stop data collection time point within the operating room.

Post Intervention: Multiple lab draws at various time points to assess sufentanil concentrations and pharmacokinetics, we will follow pain scores and opioid requirements for 24 hours post intervention, we will follow the patient until discharge from the hospital for ICU length of stay and hospital length of stay. No further follow up after discharge. We anticipate a 24-48 hour ICU length of stay and a 5-7 day hospital length of stay.

##### Sample Size and Population

Adults between ages 19 and 80 with reasonable cardiac and pulmonary function presenting for routine cardiac surgery including coronary artery bypass graft and aortic valve replacement who qualify for “fast track extubation”. Based on estimates for time to extubation for our study population on sufentanil bolus dosing where we have an average time to extubation of 190.5 minutes (SD = 99 min) and assuming 80% power and statistical significance alpha level of 0.05, our power analysis suggests that we would need to recruit at least 43 subjects into each intervention group for a total of 86 subjects giving the ability to detect a 60 min difference in time to extubation between groups. We plan on inflating this number to at least 100 subjects with approximately 50 subjects in each arm to allow for potential minimal losses in evaluable patients associated with unforeseen attrition related to exclusion criteria and censoring due to death or severely poor clinical state, but may enroll up to 150 subjects.

#### STUDY TEAM ROSTER

**Lead Investigator:** Jeffrey Songster, MD

*984455 Nebraska Medical Center*
*Omaha, Ne 68198-4455*
*308-520-2847 cell*
**
*Document delegation of responsibilities to research staff, oversee the conduct of the trial and ensure participant safety and conduct in accordance with the protocol and good clinical practice*

**Mentor/Co-Investigator:** Nicholas Markin, MD

*984455 Nebraska Medical Center*
*Omaha, Ne 68198-4455*
*402-559-4081 work*
**
*Provide mentorship to the lead investigator, help ensure safe and proper conduct of the study*

**Research Staff:** Lace Sindt, RN

*984455 Nebraska Medical Center*
*Omaha, Ne 68198-4455*
*402-559-2905 work*
**
*Coordinate IRB, participant inclusion and consent, and data collection with the lead investigator*

#### PARTICIPATING STUDY SITES

#### UNIVERSITY OF NEBRASKA MEDICAL CENTER

PI: Jeffrey Songster, MD
4400 Emile St. Omaha, NE 68198
402-559-7370


### 1 STUDY OBJECTIVES

##### Hypothesis

We hypothesize that routine cardiac surgery patients receiving sufentanil infusion will have a lower sufentanil concentration at the end of surgery and have a shorter time to extubation than patients receiving sufentanil bolus dosing even with similar total doses of sufentanil.

#### 1.1 Primary Objective

We will determine if sufentanil infusion administration intra-operatively during routine cardiac surgery will decrease time to extubation from OR stop data collection time point by 60 minutes compared to sufentanil bolus administration.

#### 1.2 Secondary Objectives

We will determine if sufentanil infusion administration intra-operatively during routine cardiac surgery will result in a lower plasma concentration of sufentanil at time of arrival to ICU and if the plasma concentration correlates with time to extubation. We will determine if there is infusion vs bolus results in a difference in post-operative pain scores in the first 24 hours post-operatively, opioid requirements in morphine equivalents in the first 24 hours post-operatively, vasopressor and inotrope usage with the OR stay, early rate of reintubation in the ICU, ICU length of stay, and hospital length of stay.

### 2 BACKGROUND AND RATIONALE

#### 2.1 Background and Study Rationale

Opioid use disorder has reached epidemic levels in the United States. Based on the 2015 National Survey on Drug Use and Health, 91.8 million Americans used prescription opioids, 11.5 million misused them, and 1.9 million had opioid use disorder [1]. Cardiac surgery is often associated with high dose opioid use in the perioperative period. A single center study reported in 2019 an incidence of chronic opioid use greater than 90 days following coronary artery bypass surgery of 3.2% in opioid naïve patients and 21.7% in previously opioid exposed patients [2]. Although there are positive benefits such as analgesia, decrease sympathetic response, and myocardial protection; opioids in cardiac surgery also have side effects including: delayed emergence from anesthesia, prolonged mechanical ventilation, nausea, hyperalgesia, opioid use disorder, hypotension, vasopressor use, suppressed respiratory drive including apnea, constipation, and ileus.

Identified risk factors for prolonged mechanical ventilation after cardiac surgery are vast and include: left ventricular ejection fraction (EF) less than or equal to 30%, moderate or severe right ventricular dysfunction, moderate pulmonary dysfunction on home oxygen and/or daily bronchodilator therapy, end stage renal disease on hemodialysis, chronic kidney disease with GFR <30, redo sternotomy, emergency surgery, greater than 4 units of RBCs or FFP transfusion, mechanical circulatory support post-operatively [3,4,5]. Prolonged mechanical ventilation is associated with numerous complications such as delirium, increased need for renal replacement therapy, hospital cost, and mortality [6]. Changes in perioperative opioids have been safe, effective, and instrumental in the development of so called “fast track” cardiac surgery starting in the 1980s with extubation in the first six hours post operatively in the intensive care unit (ICU) [7]. Studies using target controlled infusions of sufentanil at varying concentrations for cardiac surgery, have confirmed faster times to extubation, but also showed lower rates of hyperalgesia, improved pain control, and less post-operative opioid consumption, with lower target concentrations of sufentanil 0.4ng/ml vs 0.8 ng/ml [8,9].

A common practice in cardiac anesthesia is to bolus opioids at the discretion of the anesthesiologist which could lead to peak and trough concentration variance with the potential for decrease efficacy or increased side effects when outside the therapeutic range. A study comparing continuous infusion vs. bolus administration of high dose sufentanil and midazolam for mitral valve surgery showed the bolus group required more additional doses during periods of stimulation, with increase variability of plasma concentration during the cardiopulmonary bypass period, but similar heart rates and blood pressure results [10]. The differences in plasma concentration between intermittent bolus dosing and continuous infusion are affected by total dose and timing, but may also be due to pharmacokinetic differences between the modes of administration and these differences may prove to be responsible for differences in emergence from anesthesia, analgesia, time to extubation, and other important clinical outcomes involving sufentanil. An earlier extubation along with improved analgesia and reduced hyperalgesia would improve the care and reduce the cost for cardiac surgical patients.

#### 3 STUDY DESIGN

- This trial is a single blinded randomized head to head drug administration trial comparing sufentanil administration by infusion vs bolus.
- Study groups will be balance with respect to age using a stratified randomization procedure because age is known to have an effect on time to extubation after cardiac surgery. The age strata for randomization will be age less than 60, 60-69, and 70+
- This study will take place at an inpatient academic tertiary care center in the mid-western United States of America
- The population is adult routine cardiac surgical patients presenting for coronary artery bypass grafting (CABG), aortic valve replacement (AVR), and combined CABG and AVR.
- The trial will have two arms or groups, a sufentanil infusion group and a sufentanil bolus group.
- The primary outcome is the difference in time to extubation between groups
- Based on power analysis of one month of historical data we will enroll approximately 86 patients with approximately 43 in each arm which will be powered to detect a 60 min difference in time to extubation between the groups.

- Time to extubation will be measured from the stop data collection time documented in the intra-operative anesthesia record to the time of extubation as documented by the RT/RN in the CVICU.
- The secondary outcomes include plasma sufentanil concentrations, sufentanil pharmacokinetics, 24 hour post-operative pain scores, 24 hour post-operative opioid requirements measured in morphine equivalents, intra-operative vasopressor and/or inotrope usage, ICU length of stay, Hospital length of stay, and early reintubation rate within 24 hours after extubation.
- Plasma sufentanil concentrations will be measured 5 min after pre-incision bolus, at time of post heparin ACT (pre-CPB), at removal of aortic cross clamp while still on CPB, with post protamine ACT, at time of placing first sternal wire, at arrival to ICU, at time of extubation, and 1 hour post extubation. For a total of 8 lab draws per patient.
- Labs will be labeled with the number of lab, subject group assignment, subject ID number, time, collected by research personnel (initials and provider number), and transported to the research lab for processing.

  Ex: #1 Infusion Lab; Subject ID 12345; 0740 AM; collected by AB 123456
- The sufentanil pharmacokinetics will be based on analysis of the sufentanil concentrations vs time so all doses of sufentanil will be recorded with respect to amount in (mcg) and time administered in the intra-operative anesthesia record and lab draws will be labeled with the time they were drawn.
- Pain scores will be taken and recorded by the ICU nurse at time of extubation, and every 2 hours for the first 24 hours after extubation. This will be recorded in the RN flowsheet and abstracted by the research personnel.
- All opioid medications will be documented in the MAR with respect to dose and time administered by the ICU RN and then abstracted by the research personnel for the first 24 hours after extubation. These doses will be converted to morphine equivalent doses.
- Intra-operative vasopressors will be considered as phenylephrine, norepinephrine, and/or vasopressin; inotropes will be considered as ephedrine, epinephrine, dopamine, dobutamine, milrinone. All infusions of vasopressors/inotropes will be recorded in the intra-operative anesthesia record with respect to dose give and time administered. Infusions will be given an inotrope score according to the system described by Wernovsky [11] and modified to include norepinephrine at time of stop data collection.
- We will calculate the ICU length of stay in hours starting at the time of arrival to the ICU (first vital signs recorded) to the time that ICU discharge is ordered by the ICU team (order history) so as to exclude the delays in waiting for a hospital bed to open up.
- We will calculate the hospital length of stay in days with day 0 being the operative day to the day of discharge from the hospital.
- We will track reintubation rate for any reason for 24 hours after the time of extubation.

#### 4 SELECTION AND ENROLLMENT OF PARTICIPANTS

##### 4.1 Inclusion Criteria

Inclusion Criteria:

- Scheduled non-emergency cardiac surgical patients including those with planned procedures of CABG, AVR, and combined CABG and AVR
- 19 to 80 years old
- Planned pre-operative anesthesia screening visit and/or preoperative surgical clinic visit.
- Inpatient subject that is scheduled (greater than 24hrs in advance) for a non-emergency cardiac surgical case

##### 4.2 Exclusion Criteria

Exclusion Criteria:

- Sufentanil allergy
- EF less than or equal to 30%
- Moderate or severe right ventricular dysfunction,
- Moderate pulmonary dysfunction, to include patients with at home $0_2$ and/or daily bronchodilator therapy.
- End Stage Renal Disease on Dialysis
- Chronic Kidney Disease with GFR <30
- Sternotomy Re-do
- Emergency surgery
- Greater than 4 units of RBCs or FFP combined
- Mechanical circulatory support post-operatively such as ECMO, IABP, Impella
- Not eligible for rapid wean extubation protocol
- Requires infusion of sedative medication required during ICU admission
- Greater than or equal to 15 minute ICU hold within PACU
- Pregnant or breastfeeding

##### *4.3* Study Enrollment Procedures ***Describe the method for identifying and recruiting candidates for the trial.***

- Potential subjects will be approached during a pre-anesthesia clinic visit or pre-surgical clinic visit.
- All subjects will be entered on a screening and enrollment log.
- Screen Failure subjects will be entered on screening log with reason for screen failure.
- Enrolled subjects will be randomized via REDCap.

#### 5 STUDY INTERVENTIONS

##### 5.1 Interventions, Administration, and Duration

Infusion protocol:

- Induction: 0.4mcg/kg of total body weight up to a total of for induction.
- Prior to incision: 0.4mcg/kg of total body weight attempting 3 min prior to incision
    - #1 Infusion lab – collect minimum of 5 min after pre-incision bolus (Sufentanil concentration)
- Start infusion at .004 mcg/kg/min (0.25 mcg/kg/h) of total body weight immediately after the pre-incision bolus
    - #2 Infusion lab – collect at time of full heparin dose ACT check (sufentanil concentration)
    - #3 Infusion lab – collect at time of aortic cross clamp removal (sufentanil concentration)
- Reduce dose to 0.002mcg/kg/min (0.125 mcg/kg/h) of total body weight at time of separation from bypass
    - #4 Infusion lab – collect at time of post protamine ACT check (sufentanil concentration)
- Turn infusion off when first sternal wire is placed.
    - #5 Infusion lab – collect at time of first sternal wire placement (sufentanil concentration)
    - #6 Infusion lab – collect by ICU RN at time of arrival to ICU
    - #7 Infusion lab – collect by ICU RN at time of extubation
    - #8 Infusion lab – collect by ICU RN minimum of 2 hours after extubation time

Bolus protocol:

- Induction: Induction: 0.4mcg/kg of total body weight for induction
- Prior to incision: 0.4mcg/kg of total body weight attempting to administer 3 min prior to incision
    - #1 Bolus lab – collect minimum of 5 min after pre-incision bolus (Sufentanil concentration)
    - #2 Bolus lab – collect at time of full heparin dose ACT check (sufentanil concentration)
- 5 minutes after initiating CPB bypass: 0.4mcg/kg of total body weight

    - #3 Bolus lab – collect at time of aortic cross clamp removal **<u>before sufentanil bolus</u>** (sufentanil concentration)
- At removal of aortic cross clamp: 0.2mcg/kg total body weight
    - #4 Bolus lab – collect at time of post protamine ACT check (sufentanil concentration)
- After protamine but prior to arrival to ICU: Up to 0.2 mcg/kg total body weight at discretion of primary anesthesiologist.
    - #5 Bolus lab – collect at time of first sternal wire placement (sufentanil concentration)
    - #6 Bolus lab – collect by ICU RN at time of arrival to ICU
    - #7 Bolus lab – collect by ICU RN at time of extubation
    - #8 Bolus lab – collect by ICU RN minimum of 2 hours after extubation time

#### Sufentanil Study Protocol

**Infusion Group** | **Bolus Group**

- confirm consent to proceed (Infusion Group)
- confirm consent to proceed (Bolus Group)

##### Operative Room

Infusion Group:
- start data in Epic, record sufentanil doses and times

Bolus Group:
- start data in Epic, record sufentanil doses and times

##### Induction

Infusion Group:
- bolus 0.4 mcg/kg with induction agent of choice
- open study group assignment

Bolus Group:
- bolus 0.4 mcg/kg with induction agent of choice
- open study group assignment

##### Pre - Incision

Infusion Group:
- bolus 0.4 mcg/kg, attempt minimum of 3 min prior to incision then start infusion at 0.004 mcg/kg/min or 0.25 mcg/kg/hr
- #1 infusion lab: collect minimum of 5 min after bolus

Bolus Group:
- bolus 0.4 mcg/kg, attempt minimum of 3 min prior to incision
- #1 bolus lab: collect minimum of 5 min after bolus

##### Heparinization

Infusion Group:
- #2 infusion lab: collect with post heparinization ACT

Bolus Group:
- #2 bolus lab: collect with post heparinization ACT

##### CPB Start

Bolus Group:
- bolus 0.4 mcg/kg minimum of 5 min after CPB start

##### Cross Clamp Off

Infusion Group:
- #3 infusion lab: collect at time of Ao cross clamp off

Bolus Group:
- #3 bolus lab: collect at time of Ao cross clamp off (before bolus)
- bolus 0.2 mcg/kg at time of Ao cross clamp off

##### CPB End

Infusion Group:
- decrease infusion to 0.002 mcg/kg/min or 0.125 mcg/kg/hr at time of CPB end

##### Protamine

Infusion Group:
- #4 infusion lab: collect with post protamine ACT

Bolus Group:
- #4 bolus lab: collect with post protamine ACT
- during the time between protamine and arrival to the ICU, bolus up to a total of 0.2 mcg/kg at the discretion of primary anesthesiologst

##### Sternal Wires

Infusion Group:
- stop infusion at time first sternal wire is placed
- #5 infusion lab: collect at time first sternal wire placed

Bolus Group:
- #5 bolus lab: collect at time first sternal wire placed

##### Arrive in ICU

Infusion Group:
- #6 infusion lab: collect by ICU RN at arrival to ICU
- verify rapid wean protocol

Bolus Group:
- #6 bolus lab: collect by ICU RN at arrival to ICU
- verify rapid wean protocol

##### Extubation

Infusion Group:
- #7 infusion lab: collect by ICU RN at time of extubation
- record pain score at time of extubation

Bolus Group:
- #7 bolus lab: collect by ICU RN at time of extubation
- record pain score at time of extubation

##### 2 Hr Post Extubation

Infusion Group:
- #8 infusion lab: collect by ICU RN minimum of 2 hours post extubation
- record pain score every 2 hours post extubation until 24 hours post extubation

Bolus Group:
- #8 bolus lab: collect by ICU RN minimum of 2 hours post extubation
- record pain score every 2 hours post extubation until 24 hours post extubation

##### 24 Hr Post Extubation

Anesthesia Team: use total body weight for all doses; all bolus doses may be given in divided doses; you may use the induction agent of your choice; dexmedetomidine infusion is allowable if indicated per the primary anesthesiologist; ketamine is not allowable after induction; inotrope/vasopressor infusions must be accurate at time of stop data.
ICU RN: record time of arrival, time of extubation and pain scores in form provided; extubate per "ABCDEF care Bundle in the ICU" and "Adult Ventilator Extubation" protocols; record opioid medication doses and times in MAR.
Lab Label: #(1-8) study group (Infusion or Bolus) Lab; subject ID number; time; collected by initials and provider number; and palced in subject lab cooler. Example: #1 Infusion Lab; Subject ID 12345; 0740 AM; collected by AB 123456

Induction: All Patients may receive up to 5 mg of midazolam up to the primary anesthesiologist, for anxiolysis or hypnotic effect. Induction agent and dosing per primary anesthesiologist options may include Propofol, Ketamine, or Etomidate. During induction isoflurane may be used per the primary anesthesiologist, neuromuscular blocker may be selected and dosed per primary anesthesiologist.
Maintenance: inhaled isoflurane anesthetic end tidal concentration 0.6 – 1.2% Sufentanil as above, additional midazolam if BIS without EMG activity is > 60 prior to sternal closure is allowed, up to 5 mg total for case. Dexmedetomidine infusion allowed at the discretion of the primary anesthesiologist in either patient group, but discontinue infusion upon arrival to ICU.
Reversal of NMB will be administered prior to handoff to the ICU, but dose and timing up to the primary anesthesiologist
During transport to ICU intermittent bolus of propofol allowed up to 0.5 mcg/kg no doses after arrival to ICU

Data Collection:

- Doses and times of all study medications will be recorded by the EMR in the intraoperative anesthetic record.
- All study hemodynamic data will be recorded by the EMR in the intraoperative anesthetic record

**Postoperative:**
Primary end point:

- Record time of arrival to ICU
- Record time of extubation in ICU

Secondary end points:

- Include sufentanil plasma concentration in ICU labs at time or arrival to ICU
- Record visual analog pain score at time of extubation, and every 4 hours for 24 hours after arrival to ICU.
- Pain medication doses will be recorded by ICU RN in EMR in the MAR and will be totaled at a 24 hour after arrival to ICU and converted to morphine equivalents.
- ICU length of stay will be collected in days
- Hospital length of stay will be collected in days
- Hemodynamic data collected on EMR in intraoperative anesthetic record

#### 5.2 Handling of Study Interventions

Per clinical standard sufentanil will be obtained and diluted by the anesthesia team from the omnicell as is current practice 250 mcg diluted in 25 ml total volume to a concentration of 10 mcg/ml in a 30 ml BD syringe. Doses will be calculated based on total body weight and rounded to the nearest 5 mcg (1/2 ml) increments. For the infusion, after the pre-incision bolus, the syringe of sufentanil will prime Alaris syringe pump small caliber tubing and be placed on an Alaris syringe pump and programed to administer sufentanil in mg/kg/hr using total body weight and started according to the infusion protocol. After stopping the infusion and dose verified as recorded by the anesthesia record, the labeled syringe should be reconciled and wasted with the pharmacy department according to the established institutional policy.

Randomization of the study subject assignment will be done at time of consent. Study assignment will be located in REDCap and the screening/enrollment log. The study anesthesiologist will be notified of the subject's randomization on the day of surgery, which will help ensure blinding of the patient. Since the induction process is the same between both groups, the anesthesiologist being notified of the subject's assignment on the day of surgery will not alter the subject's care.

#### 5.3 Concomitant Interventions

Standard perioperative anesthesia care

##### 5.3.1 Prohibited Interventions

None

#### 5.4 Adherence Assessment

If the provider determines it is unsafe to follow the protocol then the patient will be withdrawn from the study.

#### 5.5 Description of Evaluations

##### 6.2.1 Screening Evaluation

Consenting Procedure

Research personnel will approach the subject either during their pre-anesthesia screening visit or pre-surgical clinic visit. The study will be discussed with the subject. The subject will have adequate time to read and review the informed consent and ask all questions. If the subject wishes to participate, the subject and the research personnel will sign the informed consent and a copy of the signed informed consent will be provided to the subject. Signed consents will also be scanned into the subject's electronic medical record for review.

Screening

Subjects may be screened up to 30 days prior to their surgery. Many cardiac surgery patients are scheduled within 14 days of their clinic visit giving them enough time to be screened by research personnel. A full history and physical is routinely performed prior to cardiac surgery. Additionally, other diagnostic tests such as transthoracic echocardiography and blood chemistry and blood counts are also routinely performed prior to surgery. These would be reviewed to determine if the patient met all inclusion criteria.

##### 6.2.2 Enrollment, Baseline, and/or Randomization

Enrollment

Enrollment and randomization will be completed through the REDCap system. An enrollment and randomization log will be stored on the UNMC secure drive. Direct research personnel will be aware of the subject's treatment arm on the day of enrollment, but the study anesthesiologist will not be notified of the subject's study arm until the day of surgery to ensure subject blinding has been maintained.

Baseline Assessments

- Height, weight, sex, age and body mass index
- Ejection fraction
- Serum creatinine and GFR
- Hemoglobin
- Diagnosis of sleep apnea,
- Ambulatory use of opioid medications (measured by morphine equivalent)

- STS score

Randomization

Randomization will be completed through the REDCap system. An enrollment and randomization log will be stored on the UNMC secure drive. Direct research personnel will be aware of the subject's treatment arm on the day of enrollment, but the study anesthesiologist will not be notified of the subject's study arm until the day of surgery to ensure subject blinding has been maintained.

Based on a power analysis we plan to recruit 43 patients into each study arm for a total of 86 patients.

Follow-up Visits
*Indicate treatment and follow-up visit assessments for each visit. List all measurements and procedures in bulleted format.*

We will follow pain scores and opioid requirements for 24 hours post intervention, we will follow the patient until discharge from the hospital for ICU length of stay and hospital length of stay.

*Include allowable time window in which evaluations may take place, e.g., study visits must be performed on the weeks indicated in the Schedule of Evaluations ± X days. The evaluation time window should be as narrow as technically feasible.*

*For example:*

- *Visit 3:*
  - *Vital Signs*
  - *Treatment Administration Form*
  - *Concomitant Medications*
  - *Adverse Events*

- *Visit 6:*
  - *General Physical Examination*
  - *Vital Signs*
  - *Treatment Administration Form*
  - *Concomitant Medication*
  - *Adverse Events*
- *End of Study Visit (EOS) Visit:*
  - *List each assessment to be performed at the participant's final visit.*
- *Early Withdraw Visit:*
  - *Specify evaluations needed for participants who discontinue study intervention early.*
  - *Specify potential reasons for early termination.*

All evaluations will occur during each patient's admission for cardiac surgery.

#### *6* SAFETY ASSESSMENTS

Possible Adverse events include:

- Over sedation due to high dose sufentanil administration could possibly lead to delayed extubation. Since sufentanil has an elimination half life of 164 minutes in adults these effects should resolve over the course of hours. All patients will be endotracheal intubated with a secure airway within minutes of the first dose of sufentanil and will remain intubated until they pass all extubation criteria. If more than 5 occurrences of this adverse event occur we plan to review our protocols and determine if protocol modification would alleviate this event.
- Poor pain control due to under dose of sufentanil is possible given the protocolized nature of sufentanil administration. If this occurs rescue doses of opioid can be administered as needed by the anesthesia or intensive care unit providers. If more than 5 occurrences of this adverse event occur we plan to review our protocols and determine if protocol modification would alleviate this event.

##### 6.1 Adverse Events and Serious Adverse Events

Below are definitions for adverse events (AEs) and serious adverse events (SAEs).

An **adverse event (AE)** is generally defined as any unfavorable and unintended diagnosis, symptom, sign (including an abnormal laboratory finding), syndrome or disease which either occurs during the study, having been absent at baseline, or if present at baseline, appears to worsen. Adverse events are to be recording regardless of their relationship to the study intervention.

A **serious adverse event (SAE)** is generally defined as any untoward medical occurrence that results in death, is life threatening, requires inpatient hospitalization or prolongation of existing hospitalization, results in persistent or significant disability/incapacity, or is a congenital anomaly.

Describe any laboratory values that will be collected to assess safety. Abnormal laboratory values should be defined (e.g. two times the normal limit or outside the reference range for a laboratory).

##### 6.2 Reporting Procedures

All AEs and SAEs will be captured on a source document page. All AEs and SAEs will be reported to the PI within 24 hrs. of notice of occurrence. AEs and SAEs will be reviewed by PI for severity and relationship.
Any reportable AEs will be reported to the IRB within 5 business days, and SAEs will be reported to Institutional Review Board within 24 hrs. of PI becoming aware of occurrence. All AEs and SAEs will be reviewed during DSMB meeting, regardless if they were reported to the IRB. PI will determined what follow up is needed for each event.

##### 6.3 Follow-up for Adverse Events

AEs will be followed until patient has completed study. If AE is ongoing at end of study the PI will determine how long AE will need to continue to be followed.

##### 6.4 DSMP

Members of the DSMP will include:
Jeffrey Songster, MD
Nicholas Markin, MD
Danstan Bagenda, PhD

Once 10 subjects have completed the trial, a DSMP meeting will be held. The DSMP meeting will conduct a review of the REDCap data, to ensure completeness, and will assess for futility of the study. No statistical analysis will happen during the DSMP meeting. The DSMP meeting will be to ensure the protocol has been constructed in a way that will ensure study completion.

**Interim analyses and Stopping Rules**

During the DSMP meeting, if it is determined that 3 or more subjects were unable to complete the study due to not being able to follow the protocol guidelines the study will be stopped, and the protocol will be re-evaluated.

After 50 patients have completed the study, an interim analysis will be conducted for efficacy and futility. The DSMP members will be utilized for the interim analysis. If futility is determined during the interim analysis, the study will be stopped.

#### 7 INTERVENTION DISCONTINUATION

*List criteria for discontinuing the study intervention/product (e.g., development of toxicities, study closure by institute) for a participant and methods for determining when criteria are met.*

*Determine length of time early withdraw subjects will be followed.*

*This section should also include a discussion of replacement of subjects who discontinue early, if replacement is allowed.*

*Note: It is vital to collect safety data on any subject discontinued due to an AE or SAE. In any case, every effort must be made to undertake protocol-specified safety follow-up procedures. If voluntary withdrawal occurs, the subject should be asked to continue scheduled evaluations, complete an end-of-study evaluation, and be given appropriate care under medical supervision until the symptoms of any AE resolve or the subject's condition becomes stable.*

### 8 STATISTICAL CONSIDERATIONS

#### 8.1 Sample Size and Randomization

26 patients reviewed, 24 analyzed in final analysis, 2 excluded (1 major outlier and 1 minor outlier)We be

Power Analysis Result:

| | |
|---|---|
| Mean time to extubation | 190.5 min |
| STD of time to extubation | 99 min |
| Significance (alpha) | 0.05 |
| Beta | 20% |
| Power (1-Beta) | 80% |
| Enrollment ratio | 1 to 1 |
| Detection difference in time to extubation | 60 min |
| Sample size needed (calculated number) | 86 |
| Sample size each arm (calculated number) | 43 |

9.1.2Treatment Assignment Procedures

Subject randomization group will be stored within REDCap. If it becomes necessary for the subject to become unblinded, the PI has access to the randomization group and will be able to discuss with the subject.

#### 8.2 *Provide "stopping rules" for the study. (e.g., slow accrual, high losses-to-follow-up, and poor quality control). Provide justifications for each stopping rule (discontinue patients, continue with caution, modify protocol).*

#### 8.3 Data Analyses

*Describe the statistical methods that will be used to analyze the outcomes and other study data.*

With respect to comparison between sufentanil infusion and sufentanil bolus groups for the key primary outcomes of time to extubation and concentration of sufentanil at extubation, the key analysis will involve a comparison for differences in means of these two outcomes for these two study groups tested for statistical significance at the 0.05 significance level using an unpaired students t-test. Confounding will be adjusted for different factors using multivariate regression analyses appropriate for the error distribution of the time to extubation and sufentanil concentration levels. We expect very little censoring of cases. We will nevertheless also perform time to event Kaplan-Meier based analyses (and cox regression models for confounding adjustments) to evaluate any impact of censoring, if it does occur on the results, although unlikely. Analyses of secondary outcomes - pain scores, opioid consumption in morphine equivalents, ICU length of stay, hospital length of stay, vasopressor score will be assessed using the same methods described above. Re-intubation rates will be compared using risk ratio regression methods.

Generalized Estimating Equations (GEE) regression methods will be used to compare longitudinal changes over the protocol specified time points on the outcomes indicated (assuming Gaussian or binomial link as appropriate for continuous and binary outcomes).

Visually, beyond the Kaplan-Meier curves, error bars (with means and confidence ranges) will be produced for the different protocol time points and study outcomes of interest as indicated.

Bivariate analyses indicating comparison of distribution of key variables between randomized sufentanil infusion and sufentanil bolus random mixed groups will also be produced in tabular form

### 9 DATA COLLECTION AND QUALITY ASSURANCE

#### 9.1 Data Collection and Management Forms

- Regulatory Binder
- Screening and Enrollment Log
- Subject ICF
- Subject Binder
- Source Documents
- EDC- REDCap

#### 9.2 Quality Assurance

10.3.1 Training

*Describe types and mechanisms of training of staff for the study.*

Our anesthesiologist group will not require training on administration, but will have a written protocol for reference provided to them on the day of surgery based on the group the patient is randomized to. There will be a group specific card for induction, although that is the same for each patient, and for bolus or infusion administration after induction based on the stratified randomization that occurs after induction so as to keep the patient blinded. We will train the ICU RN / or research personnel on the data collection form for recording time of arrival to ICU, time of extubation, pain scores, collecting opioid doses and converting to morphine equivalents from the chart, ICU length of stay, Hospital length of stay. We will train the RN, RT, and ICU teams on the extubation criterial for the trial to encourage equal treatment of both arms for weaning assessment readiness and extubation readiness.

We will train the ICU RN on collection, labeling, and handling of study lab samples.

We will train the laboratory personnel on processing of the lab samples to obtain, transfer, and store plasma for future analysis of plasma sufentanil concentrations.

We will perform a 12 patient interim safety, feasibility, and protocol analysis after the first 10 patients or at least 5 patients in each arm and consider changes to the protocol if warranted.

##### 10.3.2 Monitoring

After each set of 10 patients enrolled, we will evaluate adherence to protocol by >90% of expected dose of sufentanil given and 90% of doses given with in the correct time points with a 5 min window of adherence on either side of the specified time. We will check for completeness of outcomes data.
